# A hybrid simulation-based workshop improves knowledge and confidence in the management of hemorrhagic conversion of stroke among interventional neurology trainees

**DOI:** 10.1101/2024.03.25.24304877

**Authors:** Melissa B. Pergakis, Asit Misra, Fawaz Al-Mufti, Ivette Motola, Nicholas A. Morris

**Author notes:** **Correspondence to:** Melissa B. Pergakis, MD, 22 S Greene St, G7K18, Baltimore, MD 21201. Co-senior authors (Ivette Motola, Nicholas A. Morris).

## Abstract

**Objectives:** To assess the feasibility of a hybrid simulation-based workshop at a national meeting for interventional neurology trainees focused on the management of acute ischemic stroke and tissue-plasminogen activator (tPA)-related hemorrhage.

**Methods:** In this prospective, observational, hybrid simulation-based study at a fellows’ workshop at a national conference, participants were asked to manage a patient with acute ischemic stroke in the neurointerventional suite followed by thrombolytic-related hemorrhage leading to cerebral herniation during mechanical thrombectomy. We evaluated the participants’ ability to complete critical actions that were developed based on best practices and relevant American Heart Association guidelines and the Neurocritical Care Society’s Emergency Neurological Life Support protocols. The primary outcome was the improvement in knowledge from a pre-course to post-course test. Secondary outcomes included participant reactions.

**Results:** Sixty trainees completed the simulation session in 8 groups. The mean sum of critical actions completed by trainees was 9.75/14 (70%). There was a moderate effect of the intervention on trainees’ knowledge from pre-test (mean 3.8, standard deviation (SD) = 0.3) to post-test (mean 4.3, SD=0.3). The simulation scenario was described as moderately realistic, very engaging, and extremely satisfactory. Following the workshop, all fellows endorsed an increase in proficiency and confidence in neurological emergency management.

**Conclusions:** Simulation-based workshops at national conferences are feasible and a potentially useful tool for safely educating a large audience of trainees who may not have access to high-fidelity simulation platforms at their institutions.

## Introduction

Mechanical thrombectomy is the standard of care in acute ischemic stroke with anterior large vessel occlusion presenting within 6 hours of symptom onset.^1^ Additionally, the number of these procedures has increased partly due to the extended time window for mechanical thrombectomy in patients with favorable advanced perfusion imaging.^2,3^ It is suspected that these numbers will continue to rise as we find additional patient populations, including those with early ischemic changes on computed tomography imaging, that may benefit from intervention with mechanical thrombectomy.^4^

Neurointerventional procedural complications in acute ischemic stroke have been reported in up to 29% of patients including dissection, arterial perforation, vasospasm and resultant brain injury including larger ischemic territory, hemorrhage, and/or intracranial hypertension.^5^ As fellows in interventional neurology come from diverse backgrounds, some may not have experience in diagnosing and managing these events. Because of the low frequency, but high morbidity and mortality associated with these complications, it is essential that interventional neurology trainees have the opportunity to practice these complicated clinical scenarios in a supportive learning environment without exposing actual patients to harm. Simulation is an ideal learning modality to safely teach and evaluate psychomotor and nontechnical crisis resource management skills. However, neuroendovascular simulators have only recently been integrated into trainees’ clinical education.^6^ Few training programs have access to neurointerventional simulators.

To address this gap, we attempted to determine feasibility of a brief, hybrid simulation-based workshop at a national meeting, with the hope of expanding educational access among the neurointerventional training community. We were interested in the effect of the workshop on trainees’ knowledge about the diagnosis and management of hemorrhagic conversion of ischemic stroke complicated by intracranial hypertension and cerebral herniation. We also sought feedback from trainees regarding their satisfaction, emotional engagement, and perceived realism of the simulation.

## Methods

### Setting and Study Design

This was a prospective, observational hybrid simulation-based study at a trainee workshop at the Society of Vascular and Interventional Neurology annual meeting in November 2023. It was conducted by two neurointensivists (NAM and MBP), and two emergency medicine physicians (AM and IM) with expertise in acute stroke management as leaders in the American Heart Association’s Acute Stroke Life Support® course. Participants were interventional neurology trainees attending the annual meeting’s fellow workshop and included medical students, residents, and fellows. Prior to the simulation, participants were provided a QR code linked to a survey created on Qualtrics (Provo, UT, version November 2023) regarding demographics, level of training, and prior experience. They provided self-ratings of experience and competence in managing neurological emergencies, including intracerebral hemorrhage, acute stroke initial evaluation, evaluating intracranial pressure (ICP), herniation syndrome, and post-thrombolytic complications on a 5-point Likert scale from 1 (no experience) to 5 (most experienced) and 1 (not competent) to 5 (expert), respectively. They then completed a pre-test consisting of six multiple choice questions focused on risk factors, diagnosis, and management of hemorrhagic conversion of ischemic stroke complicated by intracranial hypertension. Following the survey and pre-test, participants received a 20-minute lecture on post thrombolytic-related complications, treatment of symptomatic thrombolytic-related intracerebral hemorrhage, and intracranial pressure.

We next divided the participants into eight groups of five to eight participants. Two groups participated in the simulation simultaneously, each at a separate simulation station in a large conference room. The six groups that did not initially participate explored separate content at the conference and rotated through, two groups at a time, until all groups had completed the workshop. The groups received a 10-minute pre-briefing on the simulation environment before starting the clinical scenario; they were instructed to approach the patient just as they would in a real clinical encounter. Within each group, one participant volunteered as the primary proceduralist while a second operated as the assistant. Two participants in each group acted as observers and recorded critical action items performed by the group using a checklist provided to them. The remainder of the group acted as the treatment team.

After each scenario was complete, the instructors led a debriefing session following the Debriefing with Good Judgement format to help participants reflect on their actions, ultimately leading to behavioral change.^7^ Finally, participants were provided with a QR code linking to a post-intervention survey assessing their experience in the simulation including realism, engagement, and satisfaction on a 5-point Likert scale, as well as a post-test assessing their knowledge on neurological emergencies that were tested during the simulation. The pre- and post-test questions were the same. Participants did not receive any financial compensation for their participation.

### Clinical Simulation Case and Trainee Assessment

The case and critical actions were modified from previously described work with established validity evidence to reflect a patient in the interventional neurology suite undergoing mechanical thrombectomy.^8^ The critical action checklist was modeled after the checklists in the Neurocritical Care Society’s Emergency Neurological Life Support (ENLS) protocols and American Heart Association (AHA) guidelines.^9–11^

### Simulator and Simulation Environment

SimMan Vascular manikins (Laerdal Medical; Stavanger, Norway) were used for the simulation scenario. SimMan has both neurological and physiological signs, including pupillary constriction and dilation, respiratory patterns, and seizure-like activity. The manikin can speak through an internal speaker by the physician running the simulation. Neurological assessment, including eye movements, sensory and motor exams, cerebellar functioning, and tests of neglect, could be done by asking the nurse embedded at the bedside to describe. A monitor displayed pertinent data including labs and neuroimaging. An additional bedside monitor depicted vital sign data, including telemetry, arterial blood pressure, oxygen saturation, and temperature. SimMan Vascular (Laerdal) components included an imaging monitor, imaging control panel, cine foot pedals, introducers, guide wires, micro/aspiration catheters, and stent retrievers to simulate an interventional neurovascular case. This vascular mannikin was used in conjunction with the Mentice Vascular software (Mentice AB, Gothenberg, Sweden).

The room simulated a neuro-interventional suite (Figure 1). It included all tools needed to complete the clinical scenario such as simulated medications (etomidate, rocuronium, hydralazine, labetalol, nicardipine, clevidipine, mannitol, 3% hypertonic saline, ketamine, fentanyl, and propofol), airway equipment including nasal cannulas, non-rebreather masks, bag valve masks, laryngoscopes, Bougies, and endotracheal tubes. Blood products included simulated platelets, fresh frozen plasma, prothrombin complex concentrate, cryoprecipitate, aminocaproic acid, and tranexamic acid. The nurse embedded at the bedside was available to administer any medication requested or assist in getting any equipment needed by the team during the scenario.

**Figure 1.**
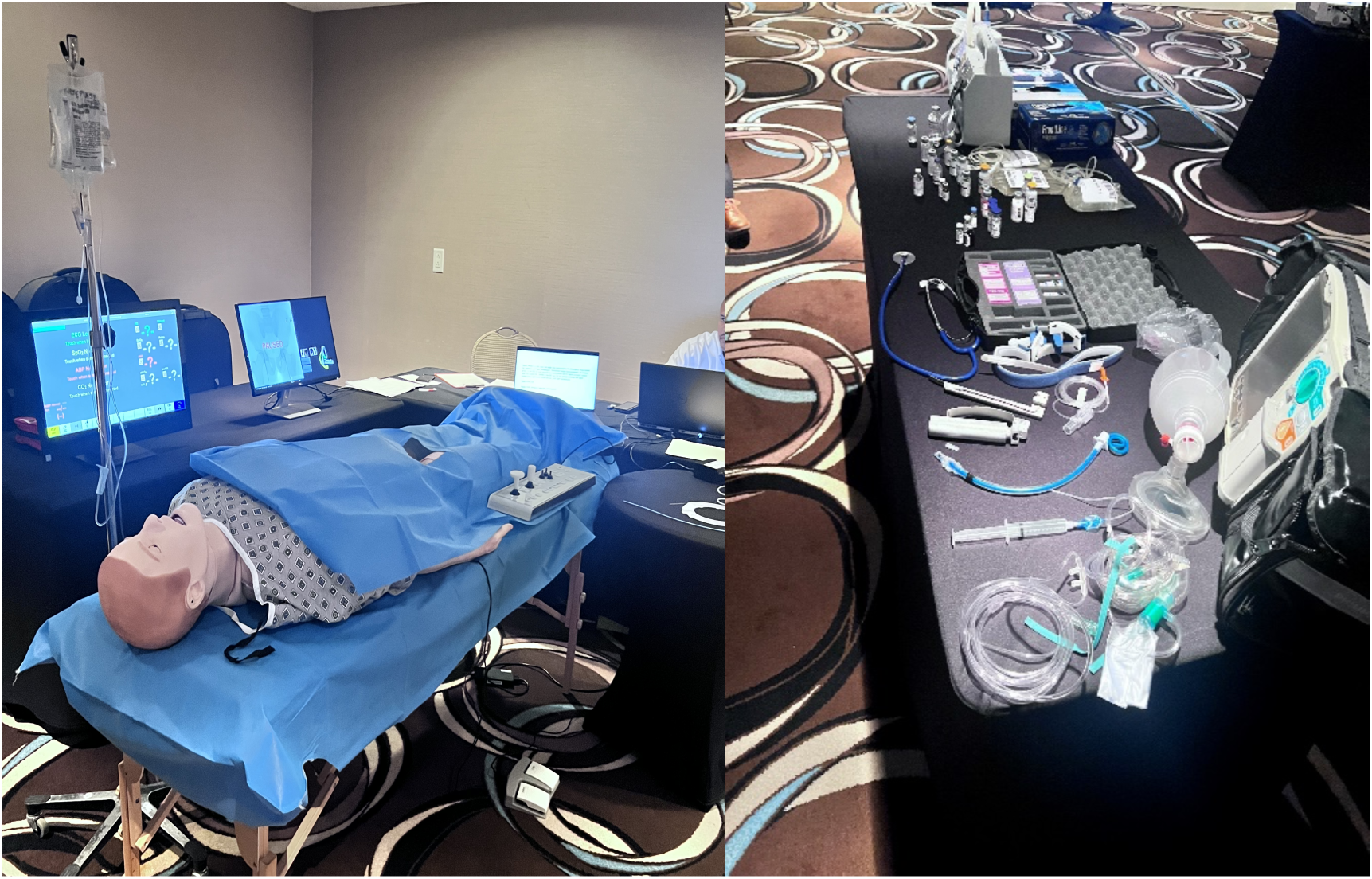
Manikin and neurointerventional suite simulation environment.

### Clinical Scenario

The clinical scenario involved a 65-year-old male with a history of hypertension and hyperlipidemia on aspirin who presented to the emergency department for aphasia and right hemiplegia. He was last known normal 45 minutes prior. In the emergency department, he was started on a nicardipine drip for blood pressure control and was given alteplase. His National Institutes of Health Stroke Scale was 22 for a full left middle cerebral artery syndrome. His finger stick blood glucose was 98 mg/dL. His computed tomography angiogram (CTA) showed a left middle cerebral artery occlusion. He was taken to the neuro-interventional suite to undergo mechanical thrombectomy. On arrival to the interventional suite, trainees were expected to perform a baseline neurological exam prior to starting the case, achieve vascular access, and attempt thrombectomy. Once trainees reached the left internal carotid artery, the patient decompensated with bradycardia, hypertension, and left-sided pupillary dilatation representing cerebral herniation syndrome. The case required trainees to recognize the change in exam, obtain STAT flat panel detector CT, stop the alteplase drip, consult anesthesia for airway management, identify post-thrombolytic hemorrhage on imaging, treat blood pressure to a goal systolic of < 180 mmHg, and reverse alteplase with cryoprecipitate, elevate the head of the bed, hyperventilate the patient, administer hypertonic saline or mannitol at appropriate doses, order appropriate labs and consult neurosurgery. (Table 2) The case ended after successful management of patient decompensation or after ten minutes had elapsed.

### Outcomes

The primary outcome was the comparison of performance on pre- and post-multiple-choice tests that assessed knowledge of the neurological emergencies. Secondary outcomes included trainee feedback regarding the perceived realism, emotional engagement, and satisfaction with the simulation case, as well as the sum score of critical action items and global rating scores ranging from one to five representing novice through expert for each group of trainees. Both the sum score of critical action items and the global rating scale were done in real-time by two independent observers (NAM and MBP) running the simulations. Themes from debriefing were tracked by instructors for qualitative review. The observers previously exhibited excellent interrater reliability.^17^

### Statistical Analysis

Descriptive statistics were reported as mean (standard deviation [SD]) for continuous variables and counts and frequencies for categorical variables. Paired two-sample *t*-tests were used to compare trainee performance on pre- and post-multiple-choice tests. Cohen’s d was used to assess effect size. Results were considered statistically significant if the p < .05. All analyses were performed using IBM SPSS Statistics 27. The reporting format is in accordance with the guidelines established by the “Strengthening the Reporting of Observational Studies in Epidemiology (STROBE)” study as well as the extended guidelines for health care simulation research.^12,13^

### Standard Protocol Approvals, Registration, and Participants Consents

This study was approved by the University of Maryland IRB, which waived the need for informed consent.

### Data Availability Statement

The data that support the findings of the present study are available from the corresponding author, MBP, upon reasonable request.

## Results

Sixty participants, mostly with a training background in Neurology, completed the simulation in 8 groups (Table 1). Participants rated baseline experience with regards to intracerebral hemorrhage, acute stroke initial evaluation, elevated ICP and herniation syndrome, and post-thrombolytic complications (Figure 2). Participants rated their competence in all of the aforementioned areas of expertise both before and after the hybrid simulation experience (Figure 3).

**Table 1.** Characteristics of Participants, n (%) unless otherwise specified.

|  |  |
| --- | --- |
| **Age** |  |
| 18-24 years old | 1 (2) |
| 25-34 years old | 50 (83) |
| 35-44 years old | 9 (15) |
| **Sex** |  |
| Female | 14 (23) |
| Male | 45 (75) |
| Prefer not to say | 1 (2) |
| **Training background** |  |
| Neurology | 54 (90) |
| Neurosurgery | 1 (2) |
| Radiology | 1 (2) |
| Other | 4 (7) |
| **Subspecialty training beyond interventional neurology** | 20 (33) |
| Vascular neurology | 7 (35) |
| Neurocritical care | 4 (20) |
| Did not report specific subspecialty training | 9 (45) |
| **Board certified** | 18 (30) |
| **Trained using medical simulation** | 28 (47) |
| **Completed clinical rotations in neurocritical care** | 50 (83) |
| **Basic Life Support certification** | 54 (44) |
| **Advanced Cardiovascular Life Support certification** | 54 (44) |
| **Advanced Stroke Life Support certification** | 1 (1) |
| **Emergency Neurological Life Support certification** | 10 (8) |

**Figure 2.**
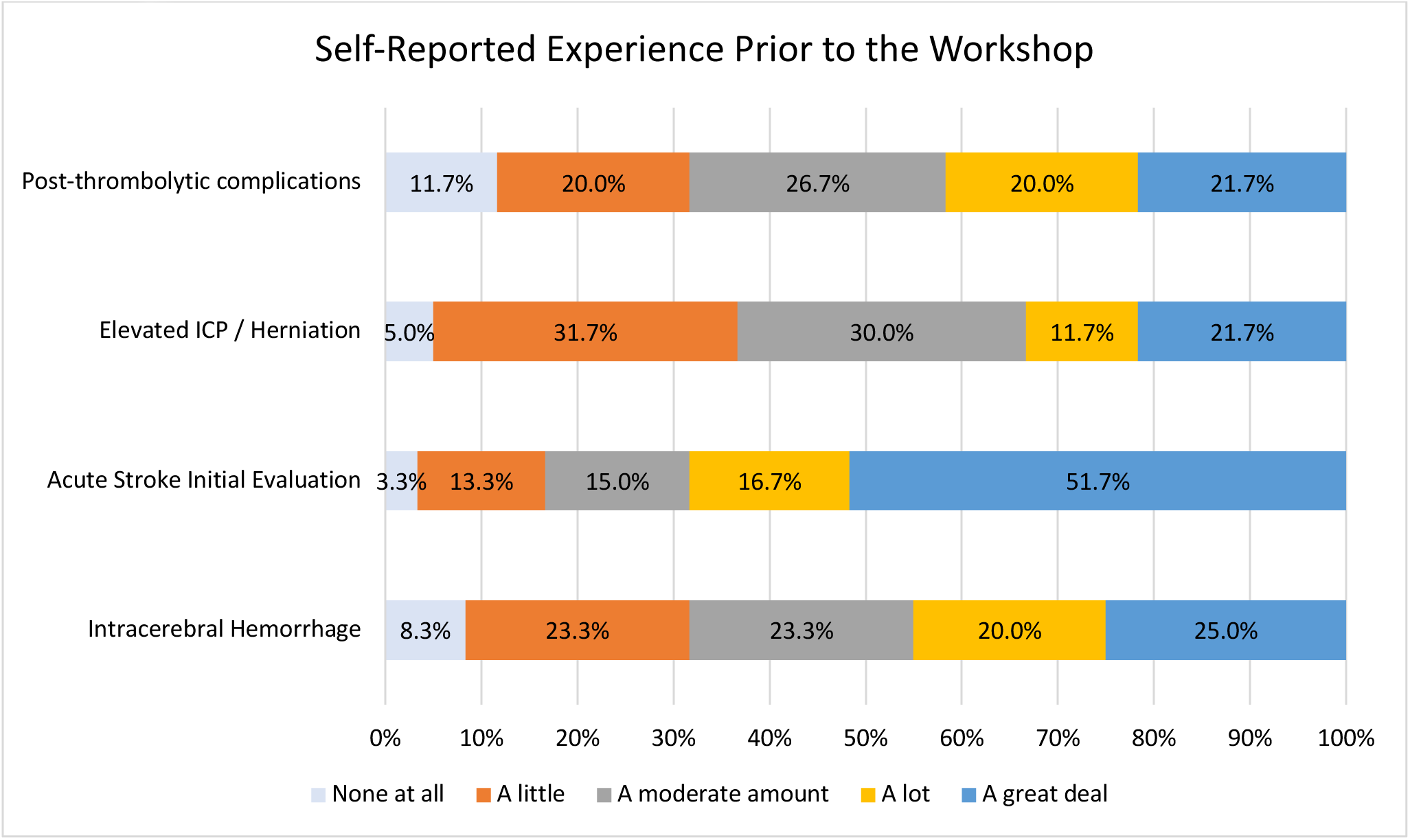
Self-reported experience prior to the workshop using a 5-point scale from *None at all to A great deal*.

**Figure 3.**
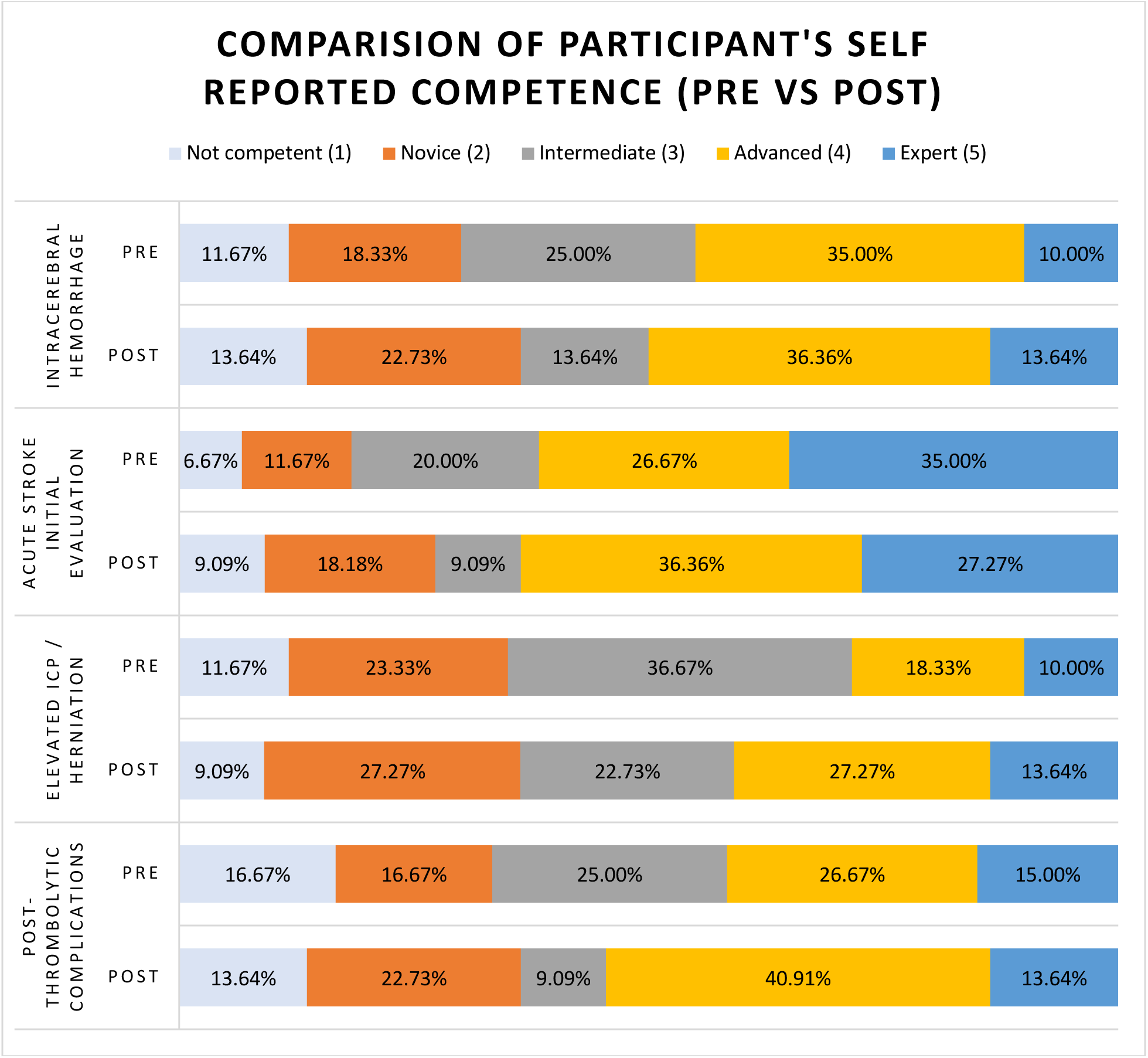
Comparison of Self-reported competence before and after the workshop using a 5-point scale from *Not competent to Expert*.

The mean sum of critical actions completed by trainee teams was 9.75 out of a total of 14 critical actions (70%; SD = 2.31) (Table 2). The mean global rating scale score for trainee teams was 3 out of 5 (SD = 0.93). All eight teams obtained flat-panel detector CT imaging when a change in exam was noted and recognized post-thrombolytic hemorrhage on imaging. Eighty-eight percent of teams consulted anesthesiology for an airway at the appropriate time, treated blood pressure appropriately after hemorrhage was identified, and treated intracranial hypertension with hypertonic saline or mannitol. Only 3/8 teams (38%) performed a baseline neurological examination at the start of the case and properly dosed hypertonic saline or mannitol. Labs including fibrinogen and coagulation factors were only ordered by 2/8 teams (25%).

**Table 2.** Critical Action, n (%)

| **Critical Action, n (%)** |  |
| --- | --- |
| Critical Action | Number of groups correctly performed<br>N (%) |
| Perform neurological examination at the start of the case including brainstem reflexes and motor exam | 3 (38) |
| Recognize herniation event by noticing pupillary dilatation | 6 (75) |
| Stop tissue plasminogen activator once change in exam is noted | 5 (63) |
| Obtain flat panel detector CT when change in exam is noted | 8 (100) |
| Consult anesthesia or airway team for intubation when appropriate | 7 (88) |
| Identify post-thrombolytic hemorrhage on imaging | 8 (100) |
| Treat elevated blood pressure to a systolic goal of at least < 180 mmHg | 7 (88) |
| Reverse thrombolytic with cryoprecipitate +/- anti-fibrinolytic | 6 (75) |
| Elevate the head of the bed | 6 (75) |
| Hyperventilate to a goal PCO<sub>2</sub> of 30-35 mmHg | 4 (50) |
| Treat intracranial hypertension with hypertonic saline or mannitol | 7 (88) |
| Properly dose hypertonic saline (30cc 23.4% or 250-500cc 3% saline, 0.5-1g/kg mannitol) | 3 (38) |
| Obtain STAT labs (must include fibrinogen and coagulation factors) | 2 (25) |
| Consult neurosurgery | 6 (75) |

Several themes emerged during the debriefing that focused on nontechnical skills. All groups showed excellent situational awareness, monitoring the status of the patient, gauging the availability of resources, and responding appropriately. Problem-solving was also recognized as quite good. Groups generally made the correct diagnosis for neurological deterioration while considering alternatives and implemented interventions quickly. Opportunities for improvement included communication skills and resource utilization. Closed-loop communication was rarely employed, and tasks were often assigned to the embedded nurse overwhelmingly without prioritization.

Twenty two of 60 (37%) participants completed the post-survey and post-test. Performance on the post-intervention knowledge assessment improved after the lecture and simulation scenario with debriefing (mean score post-test [standard deviation (SD)] = 4.3 (0.3) vs. mean score pre-test (SD) = 3.8 (0.3), p = .01) (Figure 4). The effect size was moderate, Cohen’s d = 0.44.

**Figure 4.**
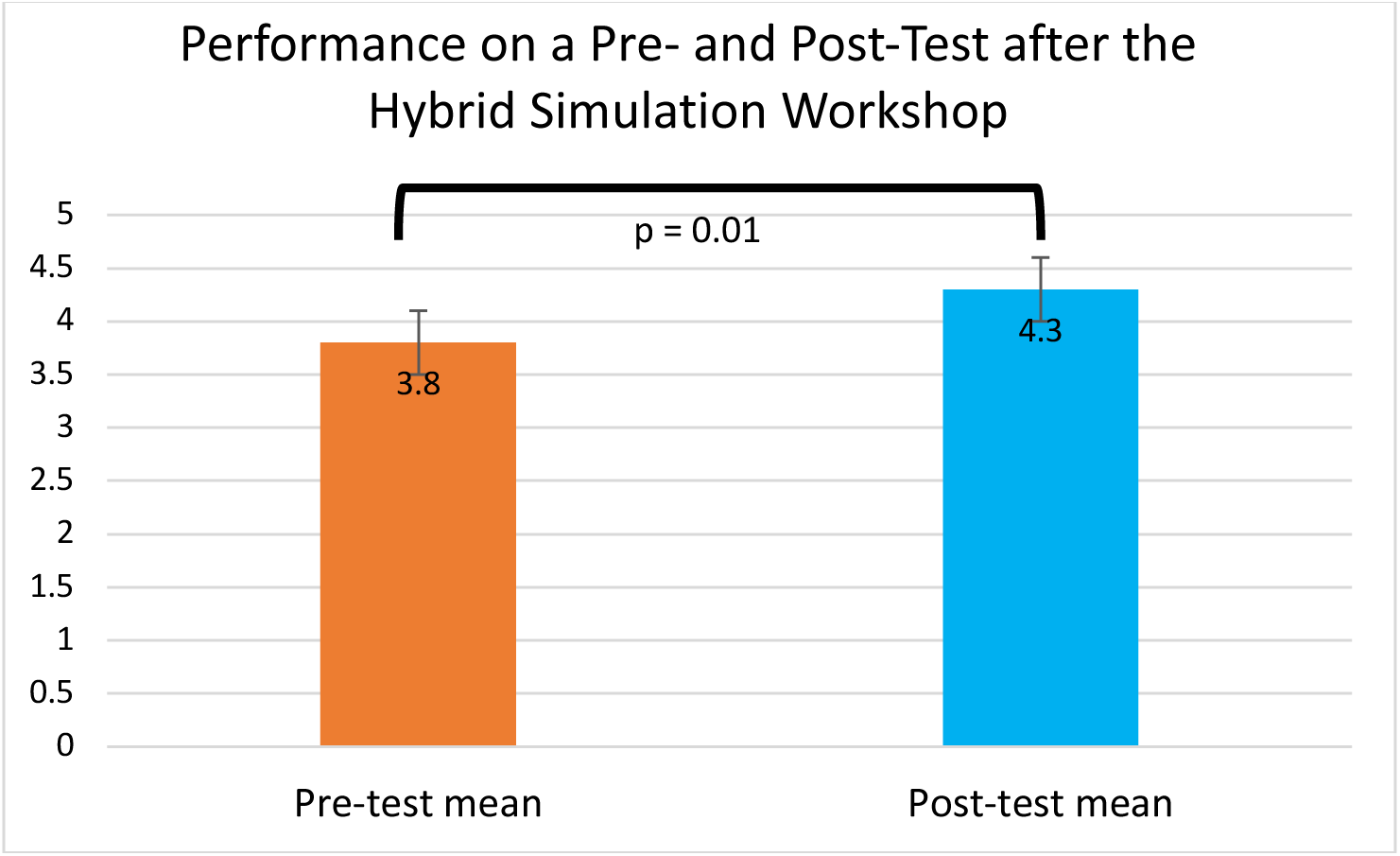
Effect of the intervention on pre- and post-test performance (mean score post-test [standard deviation (SD)] = 4.3 (0.3) vs. mean score pre-test (SD) = 3.8 (0.3), p = .01) out of 6.

Overall, participants rated the simulation scenario as moderately realistic (mean score [standard deviation (SD)] = 2.9 (0.68), range 1 (not at all realistic) to 5 (extremely realistic)), emotionally very engaging (mean score (SD) = 4.23 (0.79), range 1 (not at all engaging) to 5 (extremely engaging)), and very satisfying (mean score (SD) = 4.59 (0.49), range 1 (not at all satisfied) to 5 (extremely satisfied)). All the respondents (n=22) either somewhat agreed or strongly agreed that participation in this simulation-based workshop benefited their education/practice, helped increase their confidence in managing neurological emergencies, and helped increase their proficiency and experience with neurocritical care and emergency neurology. Also, 19/22 (87%) of respondents somewhat agreed or strongly agreed that they would prefer simulation-based training over didactic lectures to learn neurological emergencies (Figure 5).

**Figure 5.**
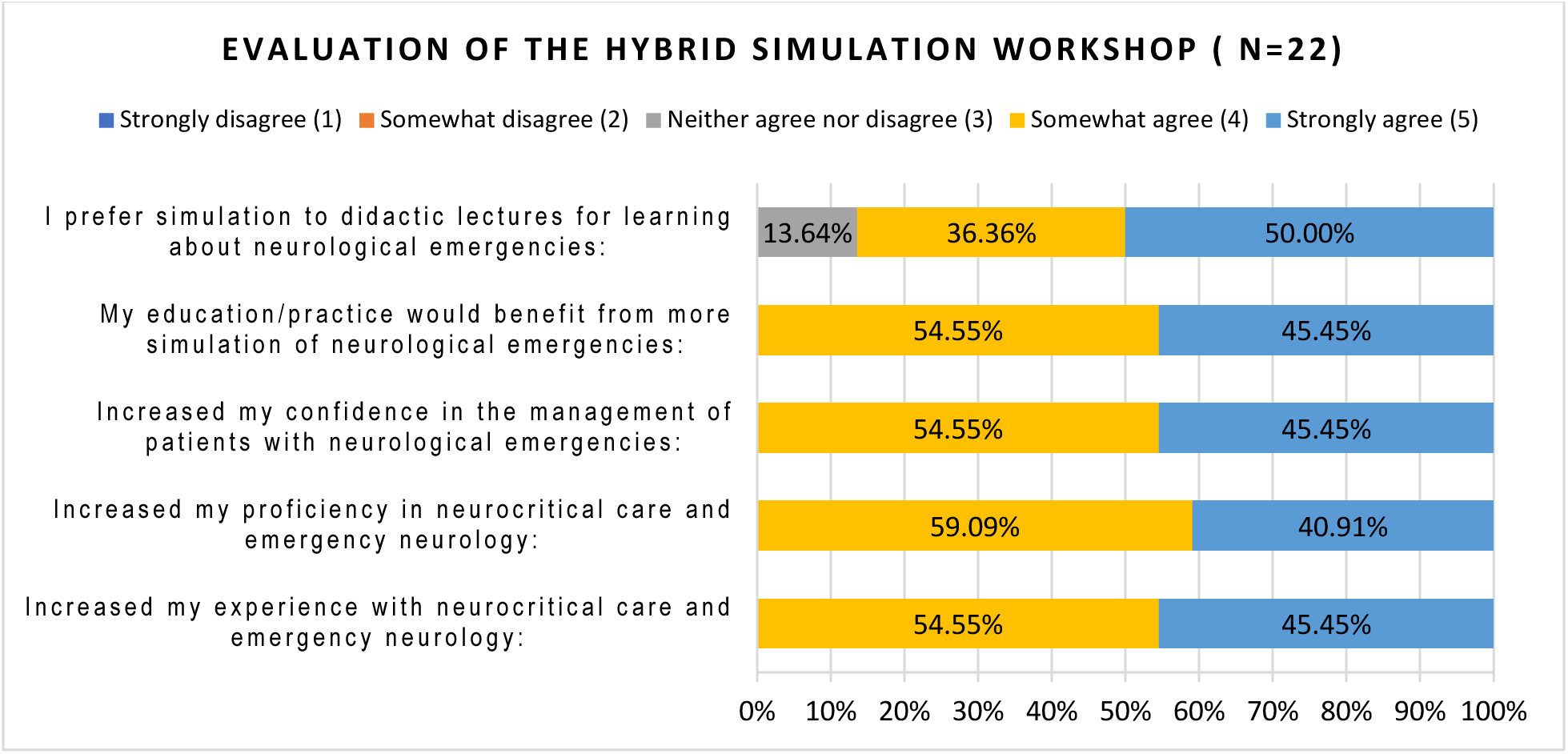
Evaluation of the simulation workshop using a 5-point scale from *Strongly disagree* to *Strongly agree*.

## Discussion

We demonstrated that it is feasible to deliver a hybrid, simulation-based workshop to 60 interventional neurology trainees at a national conference, expanding access to this powerful but expensive educational modality. Participants improved their knowledge of risk factors, diagnosis, and management of hemorrhagic conversion of ischemic stroke complicated by intracranial hypertension. They reported a high degree of engagement and satisfaction with the simulation scenario, and a moderate level of perceived realism.

As the need for interventional neurologists expands, it is essential to consider how to train the growing workforce. Only recently has the field of interventional neurology adopted the use of endovascular simulators showing both construct validity as well as improved performance in beginner trainees.^6^ Our simulation scenario study, instead, focused largely on medical management of intraprocedural complications and nontechnical skills.

Regarding medical management, most groups appropriately reversed thrombolytic-related hemorrhage with cryoprecipitate, few groups sent the appropriate lab work to assess the ongoing need for blood product resuscitation in line with current guidelines. Given the high degree of morbidity and mortality associated with post-thrombolytic hemorrhage, appropriate and rapid reversal is essential.^11^ In our previous study of trainees managing a simulated case of post-thrombolytic hemorrhage, only 38% of trainees reversed thrombolysis appropriately as opposed to 75% of groups in our current study.^14^ We suspect that the improved performance was due to several differences between the studies. First, in the current study participants were primed with a short, high-yield didactic session delivered prior to the simulation scenario; a hybrid approach consisting of a traditional lecture combined with simulation has previously been shown to be effective in teaching the management of patients with intracerebral hemorrhage and neurological deterioration.^15^ Second, in this study participants managed the patient as part of a group, whereas participants worked individually in our prior study. Finally, our prior study included mostly neurology residents whereas the current study was focused on fellows. We previously demonstrated an effect of level of training on performance.^17^

Debriefing revealed communication skills and resource utilization as two areas of crisis resource management that may deserve further attention in interventional neurology training programs. We are not aware of any studies describing nontechnical educational topics across interventional neurology training programs. The addition of crisis resource management principles to hyperacute stroke care may augment emergency preparedness and foster development of high-performance teams.^16^ A systematic review concluded that crisis resource management skills learned in simulated cases can transfer to clinical settings and may improve patient outcomes.^17^

Unfortunately, trainees at many institutions may not have access to high-fidelity endovascular simulators.^18^ Our study demonstrates that it is possible to reach many trainees at an annual meeting workshop that may not otherwise have had access to these realistic simulators. A logistical limitation was that there were too many trainees to allow for a hands-on experience for all. Regardless, the group showed improved knowledge, and endorsed emotional engagement, satisfaction, and increased confidence.

Our study has several limitations. While the workshop was intended only for fellows, the demographics survey indicated that some participants were younger than would be expected. Through conversation, we learned that a minority of participants were medical students or neurology residents. In addition, as trainees specifically registered for both the conference and the workshop, they may not represent interventional neurology trainees as a whole. Indeed, demographics indicated that trainees with a clinical neurology background were overrepresented in our cohort. Our pre- and post-test assessments were very short due to logistical reasons and no validated instrument existed. Finally, despite 60 trainees completing the hybrid simulation course and the pre-test, only 22 trainees completed the post-test and post-course evaluation. We cannot know how this may have skewed the results. Future workshops may consider requiring completion of post-surveys prior to exiting the workshop.

## Conclusion

A brief, hybrid simulation-based fellows’ workshop was feasible at a national interventional neurology meeting, improving access to this powerful educational modality that increased participants’ knowledge and confidence in the diagnosis and management of intraprocedural hemorrhagic conversion of ischemic stroke complicated by cerebral herniation.

## Acknowledgement

The authors would like to thank the support of the Gordon Center for Simulation and Innovation in Medical Education at the University of Miami Miller School of Medicine for their assistance in this project.

